# Automated Classification of Clinical EEG Reports as Normal or Pathological Using Natural Language Processing: A Multi-Centre Study

**DOI:** 10.64898/2026.11.23.26368771

**Authors:** Jan Skorupski, Barbara Ostaszewska, Marian Dovgialo, Paweł Niedbalski, Jarosław Żygierewicz, Jacek Rogala

## Abstract

We evaluated automated classification of 6,829 de-identified free-text EEG reports from 33 Polish hospitals, independently rated by three experts. Three NLP models—fine-tuned PolBERT, a TF-IDF support vector classifier (SVC), and CatBoost—were assessed across seven binary and three-class tasks. Reference labels were derived from mean expert scores, majority voting or the Dawid–Skene (D–S) model, with different treatments of uncertain cases. Inter-rater agreement was moderate (Krippendorff’s α = 0.585) and below intra-rater agreement (Cohen’s κ = 0.731–0.769); D–S skill rankings differed from unweighted intra-rater κ rankings. PolBERT led on six of seven tasks (binary accuracy 0.876–0.952), although its advantage over CatBoost on three-class tasks was not significant. On Task 2A, all models exceeded all individual experts (0.762– 0.774 vs 0.689–0.726). Binary accuracy held at two isolated hospitals; on Task 2A, SVC fell from 0.762 to 0.616, while PolBERT reached 0.797. Errors clustered in disputed reports (accuracy 0.663–0.771 in intermediate strata). Exploratory zero-shot Qwen3.8-27B achieved binary accuracy of 0.829–0.929, below supervised models, and agreed with experts similarly to their mutual agreement (κ = 0.53–0.61 vs 0.47–0.63), but favoured pathological labels. Automated classification of Polish clinical EEG reports is feasible across centres, with performance comparable to individual experts. Model probabilities were well calibrated (ECE ≤ 0.035), supporting threshold-based triage. Accuracy depends on expert disagreement and the method used to construct reference labels; label aggregation should therefore be part of the task definition.

## 1. Introduction

EEG is widely used to diagnose and monitor neurological disorders. Its clinical findings are usually documented in free-text reports, despite structured systems such as SCORE [1,2]. Differences in terminology, reporting style, and diagnostic wording across clinicians and hospitals complicate automated classification and cross-centre generalisation.

Agreement on raw EEG signal interpretation varies across readers [3], including for binary normal/pathological judgements [4], and is lowest for ambiguous patterns [5,6].

Automated classification of raw EEG has advanced with the Temple University Hospital Abnormal EEG Corpus (TUAB) [7]. Earlier studies suggested accuracy could plateau near 90% because of label disagreement [8] and described saturating gains with data and model scale [9]. However, transformer models reached 95.7–96.1% on consensus-labelled TUAB data [10]. In TUAB, explicit conclusions in reports reproduce normal/abnormal labels with 99.9% accuracy using four phrases [11]. In our dataset, as is common in free-text EEG reporting, most reports describe the findings without a standardised summary verdict (unlike ACNS-style “Impression” sections or SCORE), so the overall conclusion must be inferred from the description. Label quality has also constrained multi-site classification of Polish EEG signals [12].

EEG-report NLP has mainly supplied labels for signal classifiers [10,11,13], rather than being evaluated as a clinical classification task. Direct report studies remain mostly single-centre and English-language, and more often extract particular findings rather than an overall normal/pathological conclusion [14–17].

Two gaps remain. BERT-based models [18] perform well in clinical text applications [19,20], but transformer-based normal/abnormal classification of EEG reports has been developed at single centres with at most one external site [15], and has not, to our knowledge, been evaluated across many centres. PolBERT enables this analysis in Polish [21]. Moreover, single-rater labels and majority voting offer limited treatment of ambiguous cases. The Dawid–Skene (D–S) model [22] estimates rater-specific reliability and probabilistic consensus labels.

Large language models (LLMs) can also classify text zero-shot from instructions alone [23]. Locally hosted open-weight models avoid sending clinical reports to external services, but their performance and agreement with experts on Polish EEG reports are unknown.

Using 6,829 reports from 33 Polish hospitals rated independently by three experts, we examined (1) intra- and inter-rater agreement and D–S estimates; (2) PolBERT, TF-IDF SVC and CatBoost across seven task definitions; (3) performance relative to individual experts; and (4) transfer to two hospitals excluded from development. We also explored zero-shot classification with a locally hosted open-weight LLM.

## 2. Methods

### 2.1. Data

We analysed 6,829 de-identified free-text EEG reports from ELM19 [12], covering 33 hospitals (at least 20 reports per hospital) and including patient age and sex. Identifiers were removed by the provider. Patients were 55.16% female, with mean age 46.91 years (range 16–98); reports had a median of 61 words (IQR 47–76).

Hospital median report lengths ranged from 15 (H_14) to 107 words (H_13). Most reports lacked an explicit normal/abnormal verdict, requiring interpretation of the findings. Hospital-level demographics and report statistics are in Supplementary Table A1.

Three EEG experts (each with ≥ 10 years’ diagnostic experience) independently assigned each report to epilepsy with additional pathology, epilepsy alone, normal, other pathology, or unclassifiable. These were mapped to pathological (1; all three pathology categories), normal (0), or uncertain (0.5; unclassifiable).

For intra-rater reliability, each expert re-scored 473 randomly selected reports (6.9%). Repeats were randomly interspersed over several weeks and were not identified to the experts.

Lemmatisation with spaCy [24], stop-word removal, lowercasing, and punctuation removal did not significantly improve results and were omitted.

### 2.2. Dataset Partitioning

Using anonymised hospital IDs, we created two isolated transfer hospitals, one fixed development fold and five cross-validation folds.

H_32 (228 reports) and H_33 (236) were selected before modelling for adequate site-level sample sizes and contrasting mean expert scores (0.39 and 0.69; Supplementary Table A1). Both hospitals were excluded from training, hyperparameter selection, and cross-validation.

The other 6,365 reports from 31 hospitals were stratified into six folds, preserving hospital proportions. One fold (n = 1,061) was used only for hyperparameter development and joined the training data in every iteration; the remaining five (n = 5,304) served as cross-validation test folds. For transfer testing, models were retrained on all six folds and evaluated once at H_32 and H_33.

### 2.3. Classification Task Definitions

Seven tasks varied the handling of uncertain annotations (Table 1).

**Table 1.**
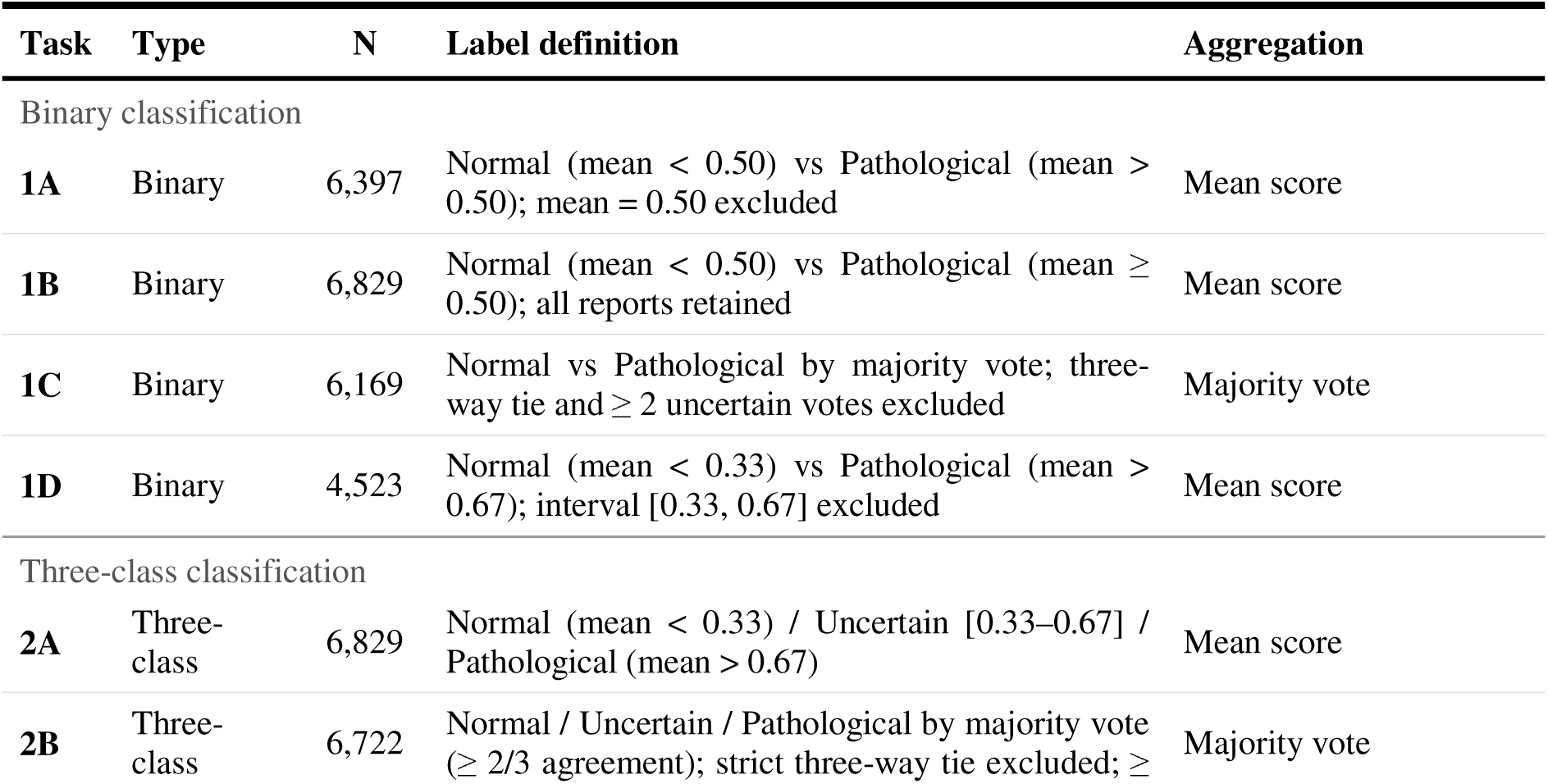
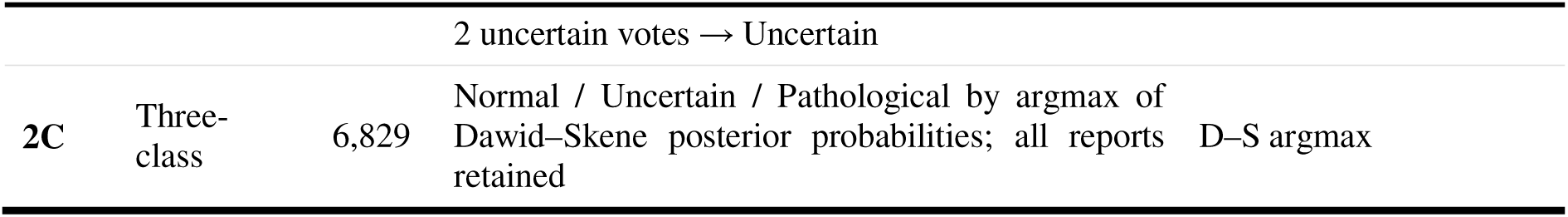
Summary of the seven classification task formulations. N = number of reports retained after task-specific exclusions. D–S, Dawid–Skene.

| Task | Type | N | Label definition | Aggregation |
|---|---|---|---|---|
| Binary classification |  |  |  |  |
| **1A** | Binary | 6,397 | Normal (mean < 0.50) vs Pathological (mean > 0.50); mean = 0.50 excluded | Mean score |
| **1B** | Binary | 6,829 | Normal (mean < 0.50) vs Pathological (mean ≥ 0.50); all reports retained | Mean score |
| **1C** | Binary | 6,169 | Normal vs Pathological by majority vote; three-way tie and ≥ 2 uncertain votes excluded | Majority vote |
| **1D** | Binary | 4,523 | Normal (mean < 0.33) vs Pathological (mean > 0.67); interval [0.33, 0.67] excluded | Mean score |
| Three-class classification |  |  |  |  |
| **2A** | Three-class | 6,829 | Normal (mean < 0.33) / Uncertain [0.33–0.67] / Pathological (mean > 0.67) | Mean score |
| **2B** | Three-class | 6,722 | Normal / Uncertain / Pathological by majority vote (≥ 2/3 agreement); strict three-way tie excluded; ≥ 2 uncertain votes → Uncertain | Majority vote |
| **2C** | Three-class | 6,829 | Normal / Uncertain / Pathological by argmax of Dawid–Skene posterior probabilities; all reports retained | D–S argmax |

Binary Tasks 1A–1D used different exclusions: 1A omitted reports with mean score 0.50; 1B included all 6,829 reports and labelled means ≥ 0.50 pathological; 1C used majority voting, excluding three-way ties or ≥ 2 uncertain votes; and 1D retained only means < 1/3 or > 2/3.

Three-class Tasks 2A–2C included an Uncertain class. Task 2A used mean-score thresholds for all reports; 2B used majority voting, accepting ≥ 2 uncertain votes and excluding only three-way ties; and 2C used the most probable class from One-Coin D–S posteriors, without exclusions.

### 2.4. Intra-rater and Inter-rater Agreement Estimation

Intra-rater agreement was measured by percent agreement, Cohen’s κ and linearly weighted κ (κω) for ordinal ratings.

Inter-rater agreement was measured by ordinal Krippendorff’s α (95% CI from 1,000 bootstrap samples), the two-way mixed-effects consistency ICC(3,1) and pairwise Cohen’s κ.

### 2.5. Dawid–Skene Expert Characterisation

We fitted the Dawid–Skene model [22] with crowd-kit v1.4 [25]. It jointly estimates each rater’s errors and posterior report labels by expectation–maximisation, treating the three categories as nominal. As no independent gold standard exists, its latent labels represent a rater-weighted panel consensus.

The primary One-Coin D–S model estimates one skill parameter s (0, 1) per rater, assuming errors are equally distributed between the other two categories. As a sensitivity analysis, the Full D–S model estimates a 3 × 3 confusion matrix for each rater, allowing asymmetric over-or under-calling of pathology.

We compared D–S P(pathological) with mean expert scores using Spearman correlation and examined reports scoring exactly 0.50. One-Coin skill rankings were compared with intra-rater rankings to distinguish agreement with the panel from self-consistency.

### 2.6. Models

We tested three classifiers on each task. PolBERT [21] used a dropout/linear classification head on pooled [CLS] embeddings, trained first with the encoder frozen (learning rate 1e-5), then jointly fine-tuned (1e-6); both stages used early stopping (patience 20 and 5, respectively) on 200 validation reports. SVC used TF-IDF features and an RBF kernel for binary tasks or a degree-four polynomial kernel for three-class tasks, with within-fold Platt calibration. CatBoost [26] used native text features (bag-of-words, BM25 and Naïve Bayes), generated within each fold, and shallower trees for binary than three-class tasks. Hyperopt Bayesian optimisation (Tree-structured Parzen Estimator) selected SVC and CatBoost hyperparameters by development-fold accuracy (Supplementary Table A3). Five-fold test performance was assessed by accuracy and macro-F1. PolBERT learning curves are shown in Supplementary Figure A1.

### 2.7. Zero-shot LLM Classification (Exploratory Analysis)

As an exploratory zero-shot classifier, we ran open-weight Qwen3.8-27B [27] (4-bit NVFP4 quantisation) through NInfer [28] on one local NVIDIA RTX 5090 GPU; no reports left the institution. Each report was processed once in reasoning mode (32,768-token budget), using the recommended sampling settings (Supplementary Text S1). Qwen3.8-27B is the highest-ranked open-weight model in the 4–160B class on the Artificial Analysis Intelligence Index v4.3.2 (34 points; ranked 1 of 227; accessed 20 October 2026) [29]; see Supplementary Text S4.

Qwen received Polish instructions and only the EEG description as input. Separate prompts produced binary labels (normal, patho; Tasks 1A–1D) and three-class labels (normal, not_sure, patho; Tasks 2A–2C), with each prediction reused across tasks of its type. Predefined rules addressed normal variants, physiological and drug effects, definite abnormalities, borderline wording, contradictions, fragmentary text, and artefacts. An EEG researcher who was not a clinical electroencephalographer drafted the prompts, consulting colleagues and LLMs; the prompts were not revised using evaluation results (Supplementary Text S1).

Experts received no operational definition of ‘unclassifiable’. The three-class prompt defined ‘not_sure’ as borderline or non-specific findings without definite abnormality, or reports too incomplete, contradictory, or artefact-dominated to interpret. The binary prompt always required a normal/pathological decision.

An invalid response triggered up to three corrective prompts (needed once). The LLM returned labels only, so calibration was not assessed.

### 2.8. Statistical Comparisons

We applied separate Friedman tests to binary (4 tasks × 5 folds; 20 blocks) and three-class results (3 × 5; 15 blocks), with Bonferroni-adjusted Conover–Iman comparisons. Shared reports and folds violate full block independence, so p-values are descriptive. Analyses used Python (scipy, scikit-posthocs, pingouin).

The zero-shot LLM was excluded from the planned Friedman tests because its predictions do not depend on training folds and are reused across tasks. Instead, pooled fold predictions were compared with each supervised model using exact McNemar tests and paired-bootstrap 95% Cis (2,000 resamples), with Holm correction for 21 comparisons. Accuracy CIs used the Wilson method; transfer-hospital comparisons were descriptive.

Task 1B calibration was evaluated across five folds with reliability diagrams, ten-bin Expected Calibration Error (ECE), and Brier score for P(pathological). SVC used within-fold Platt probabilities; CatBoost and PolBERT used softmax probabilities.

Expert accuracy was measured against each task’s reference labels (Table 6). For Tasks 1A–1C, expert uncertain votes counted as pathological (threshold ≥ 0.5); for Task 1D, they counted as errors. Three-class expert scores mapped directly to the three labels.

We compared Qwen with each expert on all 6,829 reports using binary and three-class Cohen’s κ (bootstrap 95% CIs), and recomputed Krippendorff’s α after replacing each expert with Qwen. Expert accuracy was also evaluated against the other two experts’ labels (leave-one-out). We compared uncertain-label frequencies and κ for ‘rated uncertain’, and refitted D–S with Qwen as a fourth rater for description only, not reference labelling.

### 2.9. Ethics Statement

The study used retrospectively collected reports anonymised by Elmiko Biosignals sp. z o.o. before researcher access, without identifiable individuals. Under the applicable Polish Act on the Professions of Physician and Dentist, this was not medical experimentation; ethics committee approval was not required, and individual consent was waived. Procedures followed the Declaration of Helsinki and later amendments.

## 3. Results

### 3.1. Expert Score Distributions

Mean scores were bimodal: 58.6% of reports were unanimously pathological (n = 2,155; 31.6%) or normal (n = 1,846; 27.0%; Figure 1). Hospital mean scores ranged from 0.25 (H_10) to 0.96 (H_13).

**Figure 1.**
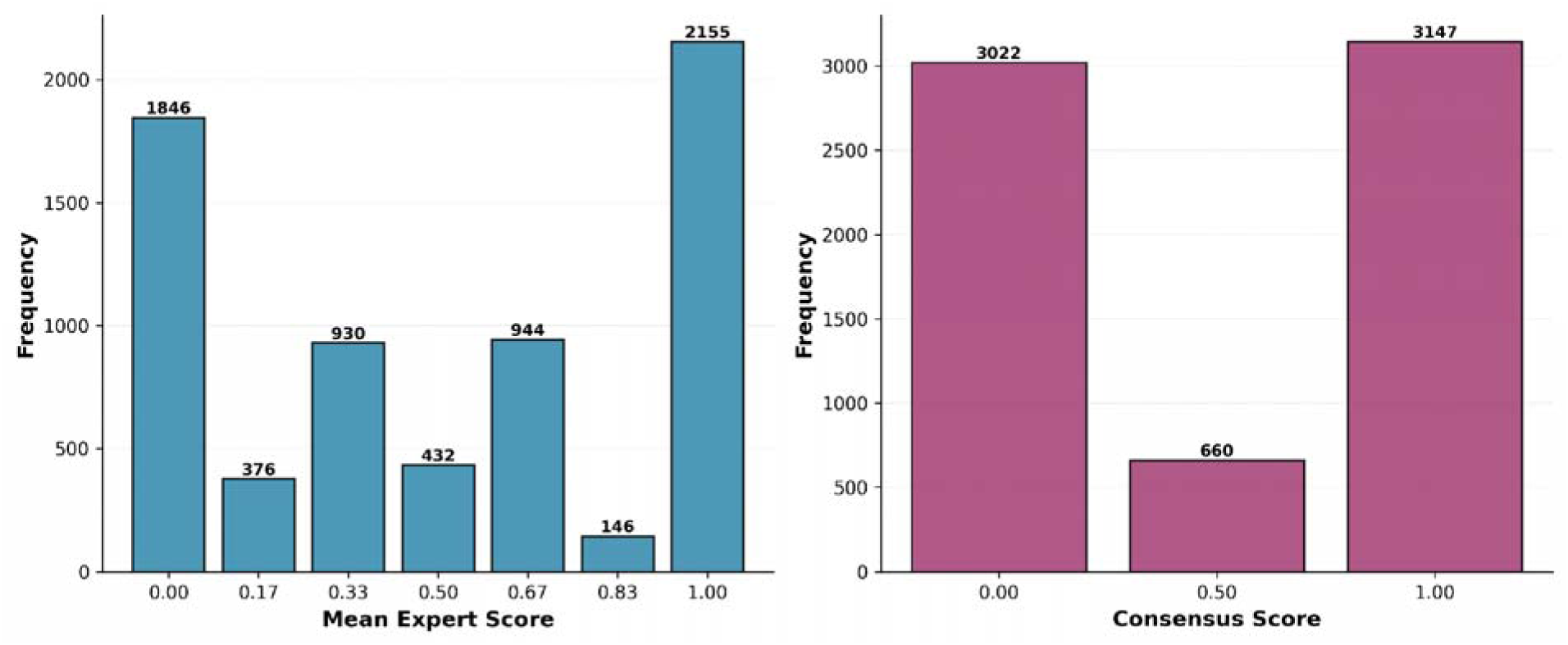
Distribution of expert scores in two variants: mean and consensus.

Majority-vote consensus was normal for 44.3% and pathological for 46.1% of reports; only 660 reports (9.7%) lacked a definite binary majority (three-way ties or ≥ 2 uncertain votes).

### 3.2. Intra-rater Reliability

Intra-rater agreement averaged 85.5% (Table 2). Only Expert B showed a substantial weighted-versus-unweighted κ difference (Δ = 0.117 vs ≤ 0.023 for A and C), indicating more non-adjacent disagreements.

**Table 2.** Intra-rater reliability metrics and Dawid–Skene skill estimates for each expert. Percent agreement, Cohen’s κ, and weighted κ were computed on 473 repeated assessments; weighted κ applies linear weights on the ordinal {0, 0.5, 1} scale. D–S skill estimates (s□) were derived from the One-Coin Dawid–Skene model fitted to all 6,829 reports. Bold indicates the highest value per column.

|  | **Intra-rater reliability** |  |  | **D–S model** |
|---|---|---|---|---|
| **Expert** | Agreement (%) | Cohen's κ | Weighted κ | D–S skill (s□) |
| **A** | 85.2 | 0.731 | 0.708 | 0.853 |
| **B** | **86.0** | **0.769** | 0.652 | 0.811 |
| **C** | 85.2 | 0.746 | **0.739** | **0.923** |

### 3.3. Inter-rater Reliability

Inter-rater agreement was moderate (Krippendorff’s α = 0.585, 95% CI [0.570, 0.600]; ICC(3,1) = 0.61, 95% CI [0.60, 0.62]). Pairwise Cohen’s κ was 0.638 (A–C), 0.597 (B–C) and 0.501 (A– B).

### 3.4. Dawid–Skene Expert Characterisation

One-Coin D–S skill was 0.923 for Expert C, 0.853 for A and 0.811 for B (Table 2). This matched the weighted κ ranking but not the unweighted κ (B > C > A). Full D–S results (Supplementary Table A2) showed that B labelled 19.3% of latent-normal reports as pathological and A labelled 21.3% of latent-pathological reports as normal; C was more balanced.

D–S posteriors correlated strongly with mean scores (Spearman ρ = 0.982, p < 0.001), but were bimodal and often pushed intermediate means (0.33–0.67) towards one class (Figure 2). Among 432 reports with mean 0.50 (107 three-way ties; 325 unanimous uncertain ratings), P(pathological) had median 0.003 and mean 0.133, favouring uncertain or normal. Thus, D–S Task 2C differs materially from mean-threshold Task 2A.

**Figure 2.**
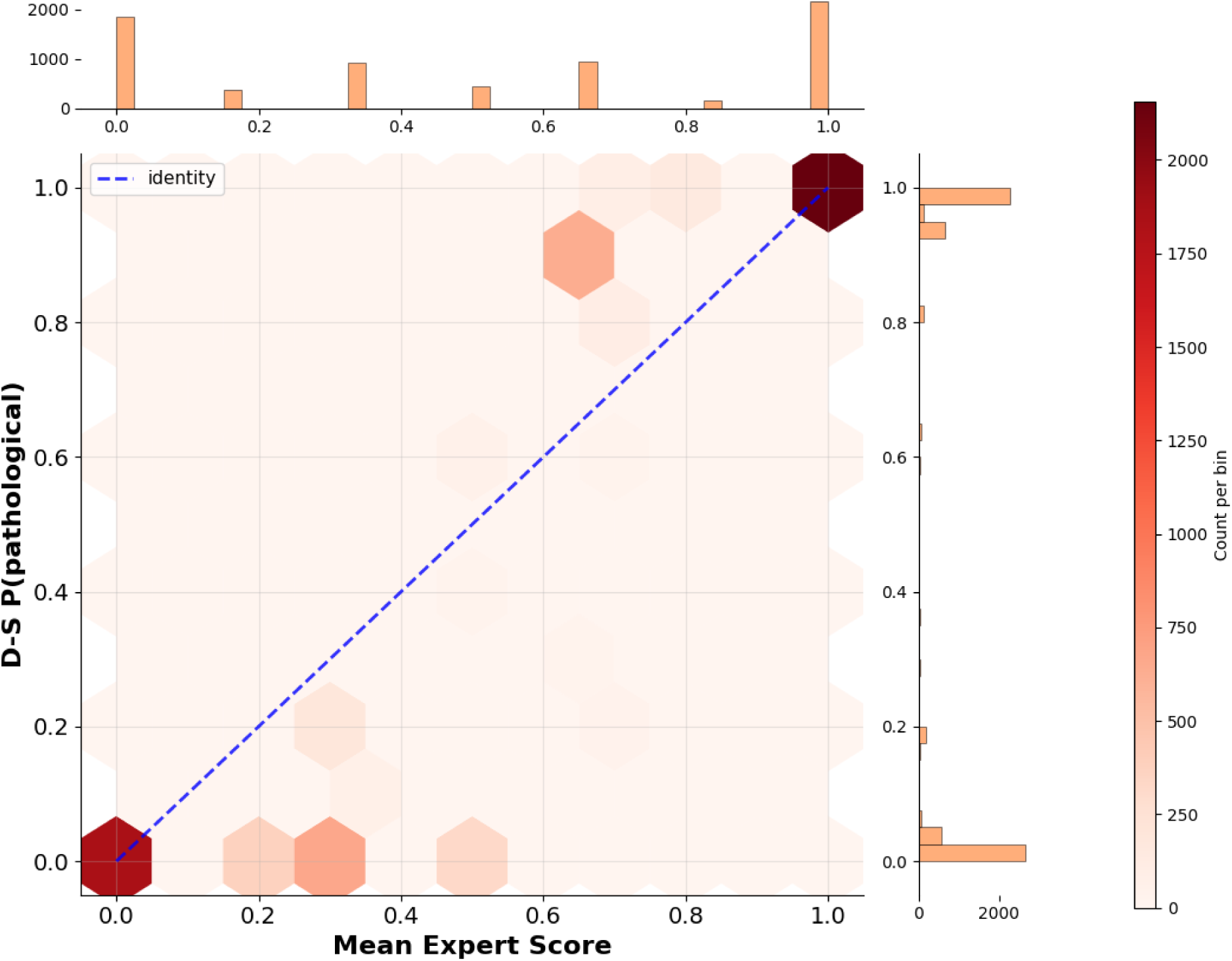
Joint distribution of Dawid–Skene posterior *P*(pathological) and arithmetic mean expert score across all 6,829 reports (Spearman ρ = 0.982, *p* < 0.001). Colour intensity indicates local point density.

### 3.5. Model Performance

Accuracy depended more on uncertain-case handling than model family (Table 3): 0.935–0.952 on Task 1D, 0.854–0.884 on Tasks 1A–1C and 0.762–0.774 on Task 2A. On Task 2B, macro-F1 (0.808–0.830) was below accuracy (0.845–0.856), reflecting the uncertain class. PolBERT led on six of seven tasks; CatBoost led on 2B. On binary tasks, SVC and CatBoost trailed PolBERT by 0.007–0.022. Across-fold SD was ≤ 0.014 for binary and up to 0.022 for three-class tasks.

**Table 3.**
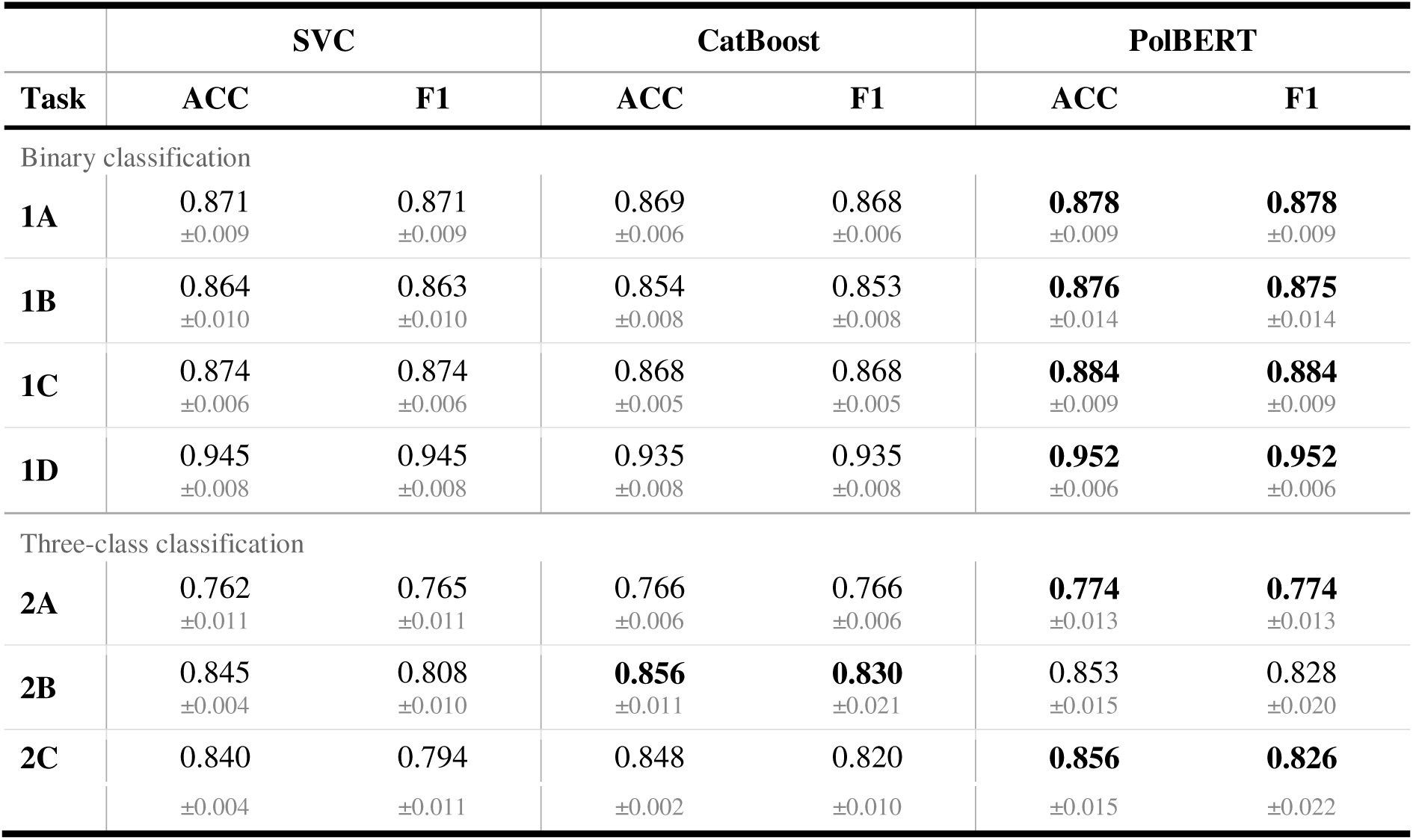
Classification performance of three NLP models across seven task formulations. Values are the mean (±SD shown below in grey) over 5-fold cross-validation. Bold indicates the best-performing model per task and metric. ACC, accuracy; macro-averaged F1 score.

|  | SVC |  | CatBoost |  | PolBERT |  |
|---|---|---|---|---|---|---|
| Task | ACC | F1 | ACC | F1 | ACC | F1 |
| Binary classification |  |  |  |  |  |  |
| **1A** | 0.871<br>±0.009 | 0.871<br>±0.009 | 0.869<br>±0.006 | 0.868<br>±0.006 | **0.878**<br>±0.009 | **0.878**<br>±0.009 |
| **1B** | 0.864<br>±0.010 | 0.863<br>±0.010 | 0.854<br>±0.008 | 0.853<br>±0.008 | **0.876**<br>±0.014 | **0.875**<br>±0.014 |
| **1C** | 0.874<br>±0.006 | 0.874<br>±0.006 | 0.868<br>±0.005 | 0.868<br>±0.005 | **0.884**<br>±0.009 | **0.884**<br>±0.009 |
| **1D** | 0.945<br>±0.008 | 0.945<br>±0.008 | 0.935<br>±0.008 | 0.935<br>±0.008 | **0.952**<br>±0.006 | **0.952**<br>±0.006 |
| Three-class classification |  |  |  |  |  |  |
| **2A** | 0.762<br>±0.011 | 0.765<br>±0.011 | 0.766<br>±0.006 | 0.766<br>±0.006 | **0.774**<br>±0.013 | **0.774**<br>±0.013 |
| **2B** | 0.845<br>±0.004 | 0.808<br>±0.010 | **0.856**<br>±0.011 | **0.830**<br>±0.021 | 0.853<br>±0.015 | 0.828<br>±0.020 |
| **2C** | 0.840 | 0.794 | 0.848 | 0.820 | **0.856** | **0.826** |
|  | ±0.004 | ±0.011 | ±0.002 | ±0.010 | ±0.015 | ±0.022 |

For binary tasks, the Friedman test found model differences (χ²(2) = 24.03, p < 0.001), all pairwise significant, with PolBERT ranked best (1.30; Table 4). For three-class tasks, differences were smaller (χ²(2) = 8.93, p = 0.011): PolBERT (rank 1.53) beat SVC but not CatBoost; CatBoost and SVC did not differ significantly.

**Table 4.** Statistical comparison of the models’ performance. Friedman test statistics and Conover–Iman post-hoc pairwise comparisons (Bonferroni-corrected). Models are sorted by mean rank within each group (1 = best); ranks are averaged over 5-fold × task blocks (n = 20 and n = 15 for binary and three-class groups, respectively). *** p < 0.001, ** p < 0.01, * p < 0.05, ns not significant.

| Model | Mean rank | vs SVC | vs CatBoost | vs PolBERT |
|---|---|---|---|---|
| Binary tasks (1A–1D)<br>χ²(2) = 24.03, p < 0.001 |  |  |  |  |
| **PolBERT** | **1.30** | * | *** | — |
| SVC | 1.88 | — | *** | * |
| CatBoost | 2.83 | *** | — | *** |
| Three-class tasks (2A–2C)<br>χ²(2) = 8.93, p = 0.011 |  |  |  |  |
| **PolBERT** | **1.53** | ** | ns | — |
| CatBoost | 1.87 | ns | — | ns |
| SVC | 2.60 | — | ns | ** |

### 3.6. Transfer Evaluation on Isolated Hospitals

At H_32 and H_33, binary accuracy was largely maintained or improved (0.841–0.961; Table 5). PolBERT led on Tasks 1B, 1D, 2A and 2B; SVC led or tied on 1A, 1C and 2C.

**Table 5.** Transfer evaluation — combined isolated hospitals. ACC, accuracy; macro-averaged F1 score. Bold indicates best-performing model per task. Red (Task 2A SVC) indicates a substantial decline relative to cross-validation performance (ACC 0.762, F1 0.765).

|  | **SVC** |  | **CatBoost** |  | **PolBERT** |  |
|---|---|---|---|---|---|---|
| **Task** | **ACC** | **F1** | **ACC** | **F1** | **ACC** | **F1** |
| Binary classification |  |  |  |  |  |  |
| **1A** | **0.908** | **0.908** | 0.887 | 0.886 | **0.908** | **0.908** |
| **1B** | 0.884 | 0.881 | 0.841 | 0.833 | **0.894** | **0.894** |
| **1C** | **0.908** | **0.907** | 0.883 | 0.881 | 0.893 | 0.892 |
| **1D** | 0.958 | 0.958 | 0.943 | 0.942 | **0.961** | **0.961** |
| Three-class classification |  |  |  |  |  |  |
| **2A** | 0.616 | 0.590 | 0.733 | 0.721 | **0.797** | **0.787** |
| **2B** | 0.845 | 0.800 | 0.816 | 0.781 | **0.850** | **0.802** |
| **2C** | **0.853** | **0.809** | 0.841 | 0.806 | 0.845 | 0.802 |

On Task 2A, PolBERT achieved accuracy/F1 of 0.797/0.787, whereas SVC fell to 0.616/0.590 from cross-validation values of 0.762/0.765, driven by H_32 (accuracy 0.474; Supplementary Table A4). On Tasks 2B/2C, all models had lower F1 at H_32 than H_33 (PolBERT 0.651/0.616 vs 0.808/0.817).

### 3.7. Comparison with Individual Expert Accuracy

Table 6 compares experts with the best model against task-specific consensus labels. Because each expert contributed to the consensus, expert accuracy measures agreement with the panel, not independent diagnostic validity.

**Table 6.** Expert accuracy against the task-specific reference label and best model accuracy. Expert accuracy is computed on all reports retained for the task (N); individual uncertain ratings were mapped as described in Methods. Best model accuracy is the accuracy on the held-out test folds of the 5-fold cross-validation (5,304 reports before task-specific exclusions). Expert values are not independent benchmarks because each expert contributed to the reference label. * On Task 1D, an expert can disagree with the reference label only through an uncertain rating, so expert values are not directly comparable with model accuracy. † Best model exceeded all three individual experts.

| **Task** | **N** | **Human experts** |  |  | **Best model** |
|---|---|---|---|---|---|
|  |  | Expert A | Expert B | Expert C |  |
| Binary classification |  |  |  |  |  |
| **1A** | 6,397 | 0.880 | 0.840 | 0.923 | 0.878 PolBERT |
| **1B** | 6,829 | 0.876 | 0.848 | 0.926 | 0.876 PolBERT |
| **1C** | 6,169 | 0.883 | 0.851 | 0.939 | 0.884 PolBERT |
| **1D**\* | 4,523 | 0.989 | 0.919 | 0.977 | 0.952 PolBERT |
| Three-class classification |  |  |  |  |  |
| **2A** | 6,829 | 0.721 | 0.689 | 0.726 | **0.774†** PolBERT |
| **2B** | 6,722 | 0.869 | 0.840 | 0.934 | 0.856 CatBoost |
| **2C** | 6,829 | 0.853 | 0.821 | 0.937 | 0.856 PolBERT |

On Tasks 1A–1C, PolBERT (0.876–0.884) exceeded B (0.840–0.851), approximated A (0.876– 0.883), and trailed C (0.923–0.939). On Task 1D, experts could err only by choosing uncertain, unlike models, which made normal/pathological errors; A (0.989) and C (0.977) exceeded the models (0.935–0.952), while B reached 0.919.

On Task 2A, all models exceeded all experts (0.762–0.774 vs 0.689–0.726). On 2B, A (0.869) and C (0.934) beat CatBoost (0.856); on 2C, PolBERT (0.856) approximated A (0.853), exceeded B (0.821), and trailed C (0.937). The best model beat B on every task, matched A within 0.002 on 1A–1C, and surpassed C only on 2A.

### 3.8. Model Accuracy as a Function of Inter-rater Agreement

On Task 1B, errors clustered at intermediate expert means (Figure 3). Accuracy was 0.935–0.963 for unanimous means 0 (n = 1,460) and 1 (n = 1,660), but only 0.663–0.696 at 1/3 (n = 742) and 0.682–0.738 at 2/3 (n = 740). At 1/2 (n = 324), labelled pathological in Task 1B, accuracy was 0.760–0.771. In the smaller 0.83 stratum (2P + 1U; n = 111), PolBERT scored 0.944 ± 0.035 versus SVC 0.802 ± 0.139 and CatBoost 0.798 ± 0.077. Thus, errors concentrated in contested cases.

**Figure 3.**
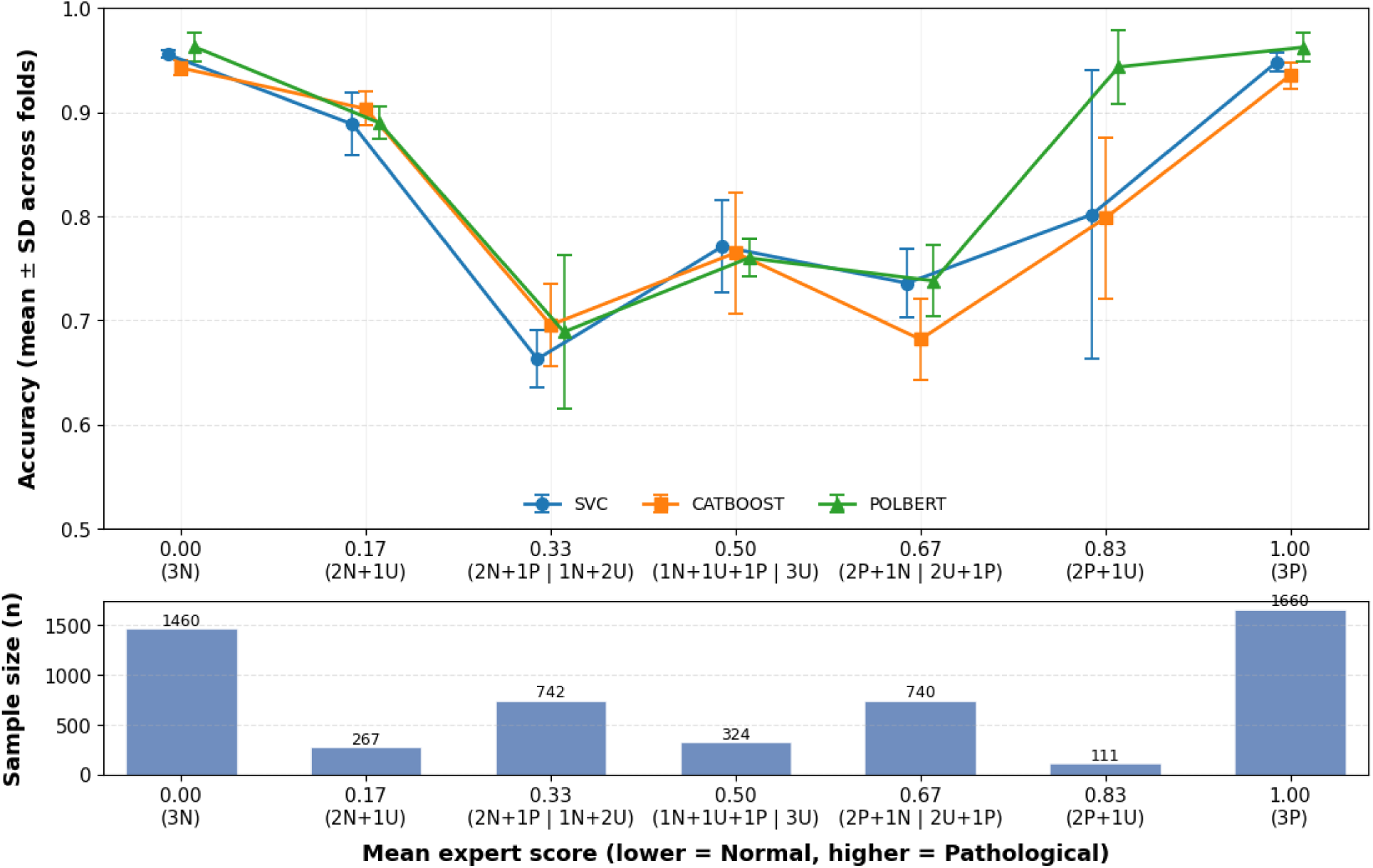
Model accuracy stratified by mean expert score on Task 1B (inclusive binary classification). Upper panel: mean accuracy ± SD across five cross-validation folds at each mean-score stratum; lower panel: stratum sample sizes in the five test folds (N = 5,304). Intermediate strata pool vote configurations: 1/3 comprises 2N + 1P and 1N + 2U, 1/2 comprises 1N + 1U + 1P and 3U (assigned to the pathological class under Task 1B), and 2/3 comprises 2P + 1N and 2U + 1P. Similar patterns were observed for Tasks 1A and 1C.

### 3.9. Model Calibration

All models were well calibrated on Task 1B (Figure 4): ECE was 0.019 for SVC, 0.032 for CatBoost, and 0.035 for PolBERT. PolBERT was more overconfident near both sides of 0.5 and more variable in sparse intermediate bins (Figure 4B), but had the best Brier score (0.090 vs 0.097 for SVC and 0.107 for CatBoost). SVC provided more faithful absolute probabilities; PolBERT had the lowest overall error.

**Figure 4.**
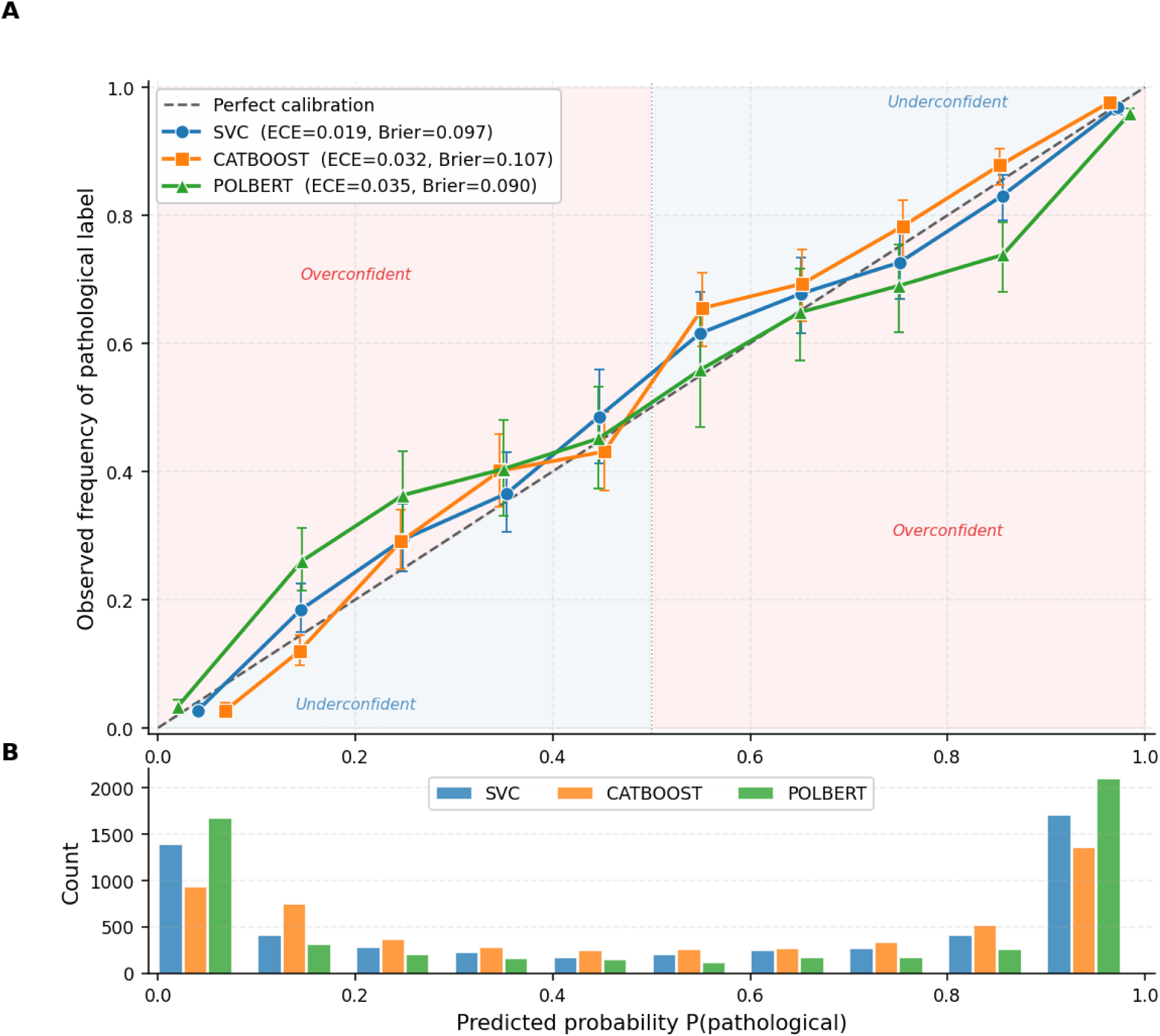
Reliability diagrams for Task 1B (inclusive binary classification, positive class: pathological). (A) Observed frequency of pathological labels as a function of predicted P(pathological); points are means across five cross-validation folds, error bars ± SD. The dashed diagonal indicates perfect calibration; shaded regions mark overconfidence (pink) and underconfidence (blue). ECE and Brier score are shown in the legend. (B) Histogram of predicted P(pathological) across all test folds; its bimodality accounts for the larger fold-to-fold variance in the intermediate bins of panel A.

### 3.10. Zero-shot LLM Reference (Exploratory Analysis)

Zero-shot Qwen achieved accuracy of 0.829–0.929 on binary tasks and 0.655–0.705 on three-class tasks (Table 7), below every supervised model. After Holm correction, 20/21 pairwise comparisons were significant (Supplementary Table A6); CatBoost on Task 1D was the exception. Gaps to the best supervised model were 0.023–0.047 (binary) and 0.119–0.153 (three-class). Three-class macro-F1 was only 0.572–0.629, reflecting weak uncertain-class performance.

**Table 7.** Zero-shot Qwen3.8-27B compared with the supervised models (exploratory analysis). Accuracy (ACC) with Wilson 95% CI on the pooled predictions of the five cross-validation test folds (N = reports retained for the task); F1 is the mean over folds, as in Table 3. Δ ACC: best supervised model minus Qwen on the same reports, with paired bootstrap 95% CI; all seven differences were significant (exact McNemar test, Holm-adjusted p < 0.001). Transfer: the two held-out hospitals (H_32, H_33).

| **Task** | **N** | **Qwen ACC [95% CI]** | **Qwen F1** | **Best supervised (ACC)** | **Δ ACC [95% CI]** | **Transfer ACC** | **Transfer F1** |
|---|---|---|---|---|---|---|---|
| ***Binary classification*** |  |  |  |  |  |  |  |
| 1A | 4,980 | 0.850 [0.840, 0.860] | 0.849 | PolBERT (0.878) | 0.029 [0.018, 0.039] | 0.875 | 0.873 |
| 1B | 5,304 | 0.829 [0.819, 0.839] | 0.827 | PolBERT (0.876) | 0.047 [0.036, 0.058] | 0.843 | 0.838 |
| 1C | 4,805 | 0.852 [0.842, 0.862] | 0.850 | PolBERT (0.884) | 0.032 [0.021, 0.043] | 0.883 | 0.879 |
| 1D | 3,498 | 0.929 [0.920, 0.937] | 0.929 | PolBERT (0.952) | 0.023 [0.013, 0.033] | 0.958 | 0.957 |
| ***Three-class classification*** |  |  |  |  |  |  |  |
| 2A | 5,304 | 0.655 [0.642, 0.668] | 0.629 | PolBERT (0.774) | 0.119 [0.104, 0.134] | 0.690 | 0.642 |
| 2B | 5,220 | 0.705 [0.692, 0.717] | 0.577 | CatBoost (0.856) | 0.151 [0.137, 0.164] | 0.706 | 0.587 |
| 2C | 5,304 | 0.703 [0.691, 0.716] | 0.572 | PolBERT (0.856) | 0.153 [0.140, 0.166] | 0.700 | 0.570 |

Qwen favoured pathology: it assigned 54.8–58.7% of binary reports to that class (reference 50.4– 53.5%) and 52.0% on Task 2A (reference 33.4%); supervised prevalence differed from the reference by at most three percentage points. On Task 1B (Figure 5), it mislabelled 11.6% of unanimously normal reports as pathological (example in Supplementary Text S2) (vs 3.7–5.8% for supervised models), but performed best for unanimously pathological reports (0.975 vs 0.936–0.963) and at mean 2/3 (0.831 vs 0.682–0.738). Accuracy was near chance at 1/3 (0.496) and 1/2 (0.512), consistent with a pathology-oriented decision threshold.

**Figure 5.**
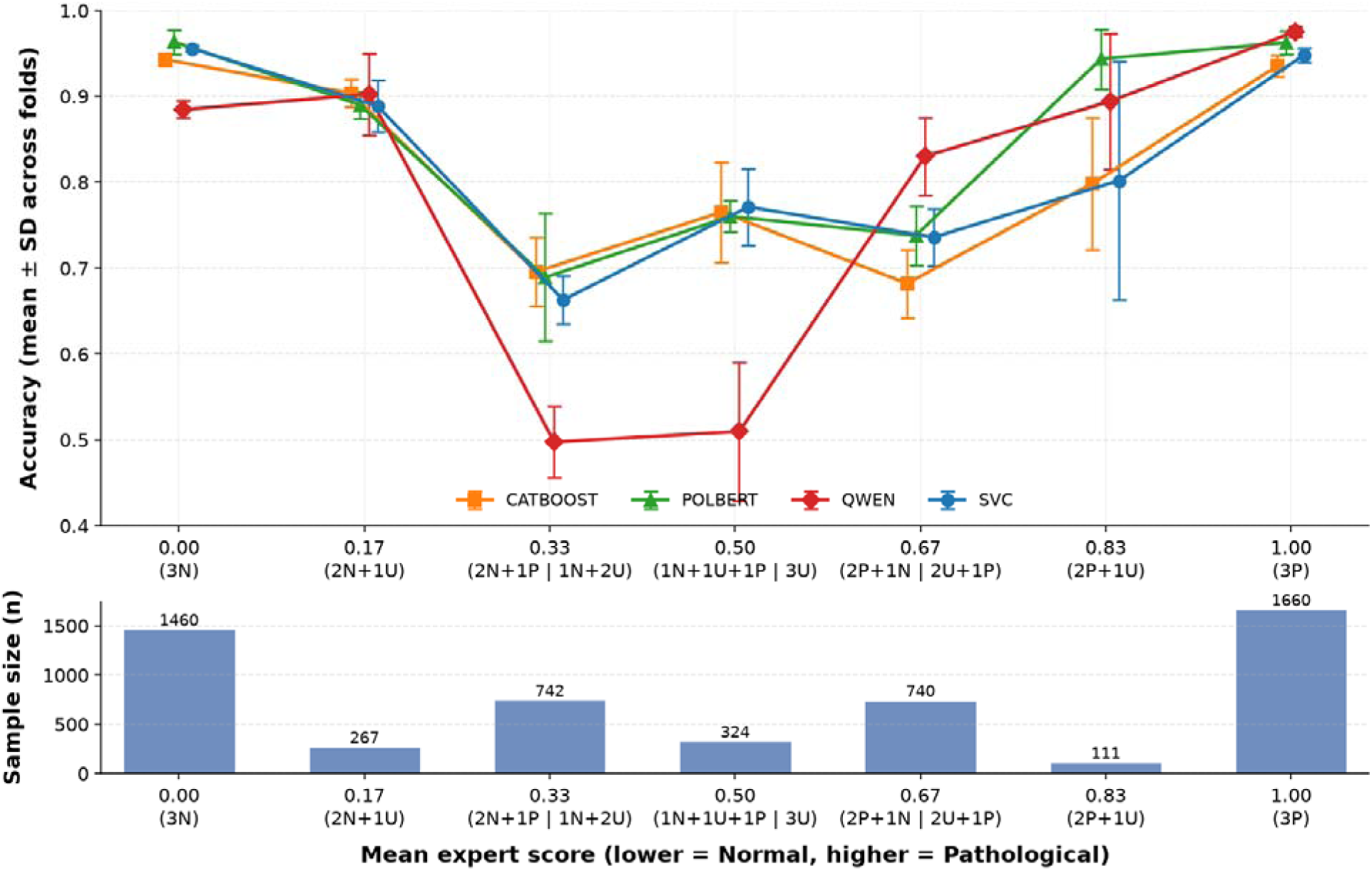
Accuracy stratified by mean expert score on Task 1B, including the zero-shot LLM (Qwen, red). Layout, strata, and sample sizes as in Figure 3; the supervised-model curves are identical to Figure 3. The corresponding plot for the three-class Task 2C is shown in Supplementary Figure A2.

Qwen used ‘not_sure’ on 17.8% of reports versus 7.3–13.5% for experts, usually on different cases: uncertain-indicator κ was 0.06–0.09 with experts versus 0.47–0.66 between experts (Supplementary Table A8). Of Qwen’s uncertain reports, 29.4% were unanimously normal to experts; 30.7% of B’s uncertain reports were normal to both colleagues. Conversely, Qwen classified 941 of Task 2A’s 1,806 reference-uncertain reports in the cross-validation test folds as pathological.

As a fourth rater, Qwen matched experts’ pairwise binary agreement (κ = 0.53–0.61 vs 0.47– 0.63; Supplementary Table A10), but had lower three-class κ (0.42–0.49 vs 0.50–0.64). Its One-Coin D–S skill was 0.737 versus 0.813–0.914 for experts. On Tasks 1A–1C, Qwen accuracy (0.829–0.852) fell within leave-one-out expert accuracy (0.754–0.888), but generally lagged on three-class tasks (Supplementary Text S3).

On H_32 and H_33, Qwen scored 0.843–0.958 on binary and 0.690–0.706 on three-class tasks (Table 7). This was not a transfer test for an untrained model; site details appear in Supplementary Table A5 and Supplementary Text S3.

## 4. Discussion

The main finding of this study is that NLP methods can classify Polish free-text EEG reports with accuracy comparable to individual experts. PolBERT achieved the highest accuracy in six of seven task formulations, but its margin was small, and its advantage over CatBoost on the three-class tasks was not statistically significant (Table 4). Classical models matched or exceeded PolBERT in several analyses: CatBoost on Task 2B, and SVC on three of seven tasks at the held-out hospitals and in calibration error. Contextual language representations are therefore valuable, but not universally preferable to simpler approaches.

Two factors are particularly important for interpreting this performance: the presence of difficult cases and systematic differences between expert raters reflected in the reference labels.

First, model performance was closely related to expert agreement. Accuracy depended much more on how borderline reports were handled than on the choice of model; the three models differed by at most 0.022 within any task (Table 3). The intermediate strata, in which experts disagreed or were undecided, accounted for more than two-thirds of all Task 1B errors for each of the three models. Thus, most model errors occurred in the same cases that were difficult for physicians to classify.

This refines the interpretation of the proposed accuracy ceiling in EEG classification [8,9]. The ceiling is not a fixed value: overall accuracy depends on the proportion of clear and ambiguous cases and on how borderline cases are defined, which complements the argument by Zhu and Western that high performance can be achieved when evaluation labels are based on expert panel consensus [10]. It also shows where label uncertainty arises. In TUAB, the verdict is stated explicitly by the reporting physician, and the label inherits that physician’s threshold [11]. In our data, three experienced readers of the same reports agreed only moderately (α = 0.585), consistent with previous EEG studies [3,4]. Report-derived labels can therefore carry two layers of individual judgement: that of the physician who wrote the report and that of the physician who reads it. Our design makes the second layer visible.

Second, disagreement between experts was not purely random. Intra-rater agreement was higher than inter-rater agreement for every expert, as also reported by Grant et al. [3], and the Dawid– Skene analysis identified stable differences in how individual physicians used the diagnostic categories. Expert B had the highest unweighted intra-rater κ but the lowest Dawid–Skene skill estimate and the lowest accuracy against the reference label on every task (Table 6), and over-called pathology, whereas Expert A tended to under-call it. This pattern is consistent with the “similar skill, individualised thresholds” model, in which experts interpret similar evidence but apply different decision thresholds [6].

Intra-rater consistency alone therefore does not demonstrate that an annotator provides unbiased labels: a physician may be highly consistent while consistently differing from the group. Because the latent classes are estimated from the same three raters, this difference is measured relative to the panel consensus, not to an external truth; Dawid–Skene analysis nevertheless separates self-consistency from alignment with the group consensus, which intra-rater agreement alone cannot do.

This may have implications for training clinical machine-learning models, although it was not tested here. A model trained on one physician’s labels may learn that physician’s diagnostic threshold together with the clinical content of the report, and reproduce over-or under-calling with high confidence; confidence-based filtering of automatically generated labels, as used in EEG pseudo-labelling pipelines [10,11], could not by itself remove such a bias. Our models were trained on labels aggregated across three experts, so this remains a hypothesis.

Multi-rater annotation and probabilistic aggregation offer a potential way to reduce this problem. In our data, Dawid–Skene produced near-binary posteriors for many reports with intermediate mean scores (Figure 2), showing that skill-weighted aggregation is not simply a rescaling of the arithmetic mean; the One-Coin model, however, does not capture asymmetric errors and therefore cannot correct a directional threshold. Whether Dawid–Skene-derived labels provide better training targets than majority voting or confidence filtering should be addressed in future work.

Because the differences in accuracy between model families were small, the higher computational cost of transformer models for training and inference should be weighed against marginal gains [30]. Model transparency matters in high-stakes clinical applications [31], but neither classical model used here is inherently interpretable (the SVC used non-linear kernels and CatBoost is a tree ensemble), so this advantage would apply only to linear models, which were not evaluated.

Binary-task performance was maintained at both fully held-out hospitals, in contrast to the cross-site degradation reported in parallel Polish multi-centre EEG signal classification [12]. On Task 2A at H_32, PolBERT (0.772) remained accurate where SVC fell to 0.474, possibly because contextual embeddings may be more robust when local terminology differs from the training data. However, the lower three-class F1 at H_32 was observed for all models, and H_32 had a much lower proportion of pathological reports than H_33; because macro-averaged F1 is sensitive to small classes, this probably reflects, at least partly, a site-level shift in class distribution rather than failure of a specific architecture. More held-out hospitals are needed before concluding that transformer models transfer systematically better.

The finding that all models exceeded individual experts on Task 2A also requires careful interpretation. Models were trained on the mean-score consensus against which each expert was evaluated, so an expert whose threshold differed systematically from the group was penalised even if their ratings were internally consistent. Higher model accuracy in this setting reflects the ability to reproduce the group-derived target, not superhuman diagnostic performance. Similarly, good calibration within cross-validation supports probability-based triage, but calibration at external sites remains to be established, and a model can be well calibrated to labels that themselves contain systematic physician bias.

The exploratory zero-shot analysis adds a different perspective. Without any training on our data, a locally run open-weight LLM reached binary accuracy a few percentage points below the supervised models and agreed with individual experts on the normal–pathological decision about as well as the experts agreed with one another. Its errors, however, were systematic: it called pathology more readily than the panel and applied the uncertain category to different reports than the experts did. The prompted model thus behaved like an additional rater with its own decision threshold, but one set largely by the written instructions. That threshold is explicit and adjustable, for example by revising the rules or calibrating the prompt on a development set, but the result reflects the prompt as much as the model.

The uncertain category illustrates this point. The experts received no operational definition of “unclassifiable”, and our prompt encoded one possible interpretation of it. The very low agreement between Qwen and the experts on which reports are uncertain (κ ≤ 0.09), together with the experts’ own differences in how often they used the category (7.3–13.5% of reports) and their only moderate mutual agreement on it (κ 0.47–0.66), suggests that part of the inter-rater disagreement may arise from an underspecified annotation category rather than from case difficulty or individual thresholds alone. Future annotation protocols should define the uncertain category operationally.

Local inference avoids transferring clinical text to external services but requires far more computation than the supervised models. The results apply to one model, one quantised build, one prompt and a single stochastic run, and the model produced labels rather than probabilities; they are a practical reference point for a locally deployable general-purpose model, not an estimate of the performance attainable with larger models or optimised prompts.

Several limitations should be noted. Experts evaluated report text without access to the raw EEG signal, so some disagreement may reflect incomplete or ambiguous reporting rather than diagnostic uncertainty. The three-level scale also collapses clinically important distinctions between different types of EEG abnormalities, which could be targets for more detailed NLP classification [14,16]. Only three annotators were available, limiting the precision of Dawid– Skene skill estimates. The same experts labelled the held-out hospitals, so transfer was tested across reporting sites but not across annotators. Because tasks share reports and folds, the statistical comparisons between models are descriptive. Finally, the study included Polish-language reports from one healthcare system and only two fully isolated hospitals; prospective validation across more centres, languages, and clinical settings is required.

## 5. Conclusion

NLP models can classify free-text EEG reports in a large, multi-centre, non-English dataset with accuracy comparable to individual experts. PolBERT showed the strongest overall performance, but its advantage was small and not consistent across tasks and sites, so computational cost should be considered alongside accuracy. A general-purpose LLM run locally without any training reached somewhat lower accuracy, with a systematic shift towards pathology, and is best regarded as an additional rater whose threshold is set by its prompt.

Achievable performance depends on the labels as much as on the models. Model accuracy falls in the same cases in which physicians disagree, so it depends strongly on case mix and on how ambiguous cases are handled, and the method of label aggregation should be reported as part of the task definition. Physician-generated labels can also carry stable individual thresholds, so they should not automatically be treated as unbiased ground truth; multi-rater annotation and methods such as Dawid–Skene can help distinguish genuine case ambiguity from systematic rater differences.

## CRediT authorship contribution statement

**Marian Dovgialo:** Writing – review & editing, Writing – original draft, Visualization, Validation, Software, Investigation, Formal analysis.

**Paweł Niedbalski:** Writing – review & editing, Data curation, Conceptualization.

**Barbara Ostaszewska:** Writing – review & editing, Writing – original draft, Visualization, Software, Methodology, Investigation, Formal analysis, Data curation.

**Jacek Rogala:** Writing – review & editing, Writing – original draft, Supervision, Resources, Project administration, Investigation, Funding acquisition, Conceptualization.

**Jan Skorupski:** Writing – review & editing, Writing – original draft, Visualization, Software, Methodology, Investigation, Formal analysis, Data curation.

**Jarosław** Ż**ygierewicz:** Writing – review & editing, Writing – original draft, Visualization, Supervision, Resources, Methodology, Investigation, Funding acquisition, Conceptualization.

## Declaration of competing interest

The authors declare that they have no known competing financial interests or personal relationships that could have appeared to influence the work reported in this paper.

## Funding

This research was funded by the ‘Center of Trustworthy AI for Life Sciences’ project carried out within the International Research Agendas programme of the Foundation for Polish Science, co-financed by the European Union under the European Funds for a Smart Economy 2021–2027 (FENG) The funder had no role in study design, data collection, analysis and interpretation, writing of the report, or the decision to submit the article for publication.

## Data availability

Data available upon request.

## Declaration of generative AI and AI-assisted technologies in the manuscript preparation process

During the preparation of this work, the authors used Claude Opus 5.5 to improve the language and readability of the manuscript, assist in identifying relevant scientific literature, and support the comparison of the methods employed in this study with the current state of the art. After using this tool, the authors reviewed and edited the content as needed and take full responsibility for the content of the published article. This statement does not concern the use of Qwen3.8-27B as a research tool, which is described in Section 2.7.

## Appendix A. Supplementary data

Supplementary material associated with this article (Tables A1–A10, Figures A1–A4 and Supplementary Texts S1–S4) is provided in a separate file.

## Notes

### Competing Interest Statement

The authors have declared no competing interest.

